# Diagnosing missed cases of spinal muscular atrophy in genome, exome, and panel sequencing datasets

**DOI:** 10.1101/2024.02.11.24302646

**Authors:** Ben Weisburd, Rakshya Sharma, Villem Pata, Tiia Reimand, Vijay S. Ganesh, Christina Austin-Tse, Ikeoluwa Osei-Owusu, Emily O’Heir, Melanie O’Leary, Lynn Pais, Seth A. Stafki, Audrey L. Daugherty, Chiara Folland, Stojan Perić, Nagia Fahmy, Bjarne Udd, Magda Horakova, Anna Łusakowska, Rajanna Manoj, Atchayaram Nalini, Veronika Karcagi, Kiran Polavarapu, Hanns Lochmüller, Rita Horvath, Carsten G. Bönnemann, Sandra Donkervoort, Göknur Haliloğlu, Ozlem Herguner, Peter B. Kang, Gianina Ravenscroft, Nigel Laing, Hamish S. Scott, Ana Töpf, Volker Straub, Sander Pajusalu, Katrin Õunap, Grace Tiao, Heidi L. Rehm, Anne O’Donnell-Luria

## Abstract

Spinal muscular atrophy (SMA) is a genetic disorder that causes progressive degeneration of lower motor neurons and the subsequent loss of muscle function throughout the body. It is the second most common recessive disorder in individuals of European descent and is present in all populations. Accurate tools exist for diagnosing SMA from genome sequencing data. However, there are no publicly available tools for GRCh38-aligned data from panel or exome sequencing assays which continue to be used as first line tests for neuromuscular disorders. This deficiency creates a critical gap in our ability to diagnose SMA in large existing rare disease cohorts, as well as newly sequenced exome and panel datasets. We therefore developed and extensively validated a new tool - SMA Finder - that can diagnose SMA not only in genome, but also exome and panel sequencing samples aligned to GRCh37, GRCh38, or T2T-CHM13. It works by evaluating aligned reads that overlap the c.840 position of *SMN1* and *SMN2* in order to detect the most common molecular causes of SMA. We applied SMA Finder to 16,626 exomes and 3,911 genomes from heterogeneous rare disease cohorts sequenced at the Broad Institute Center for Mendelian Genomics as well as 1,157 exomes and 8,762 panel sequencing samples from Tartu University Hospital. SMA Finder correctly identified all 16 known SMA cases and reported nine novel diagnoses which have since been confirmed by clinical testing, with another four novel diagnoses undergoing validation. Notably, out of the 29 total SMA positive cases, 23 had an initial clinical diagnosis of muscular dystrophy, congenital myasthenic syndrome, or myopathy. This underscored the frequency with which SMA can be misdiagnosed as other neuromuscular disorders and confirmed the utility of using SMA Finder to reanalyze phenotypically diverse neuromuscular disease cohorts. Finally, we evaluated SMA Finder on 198,868 individuals that had both exome and genome sequencing data within the UK Biobank (UKBB) and found that SMA Finder’s overall false positive rate was less than 1 / 200,000 exome samples, and its positive predictive value (PPV) was 97%. We also observed 100% concordance between UKBB exome and genome calls. This analysis showed that, even though it is located within a segmental duplication, the most common causal variant for SMA can be detected with comparable accuracy to monogenic disease variants in non-repetitive regions. Additionally, the high PPV demonstrated by SMA Finder, the existence of treatment options for SMA in which early diagnosis is imperative for therapeutic benefit, as well as widespread availability of clinical confirmatory testing for SMA, warrants the addition of *SMN1* to the ACMG list of genes with reportable secondary findings after genome and exome sequencing.

## Introduction

Spinal muscular atrophy (SMA) is a rare genetic condition characterized by progressive loss of muscle function due to the death of lower motor neurons. It is one of the most common recessive disorders, particularly in individuals of European descent where it affects between 1 in 6,000-12,000,^1,2^ while in other populations it occurs at lower but appreciable rates.^3^ In 2016, the US Food and Drug Administration (FDA) approved nusinersen, the first drug to slow disease progression, followed by the approval of onasemnogene abeparvovec-xioi in 2019, and risdiplam in 2020.^4^ The American Health Resources & Services Administration (HRSA) added SMA testing to the recommended uniform newborn screening panel (RUSP) in 2018, and it is now included in all 50 states. An increasing number of countries have also included SMA within their national screening programs. However, affected individuals may remain undiagnosed if they were born before testing became available in their region, if their parents declined newborn screening, or due to false-negative test reports.

The molecular etiology of SMA is linked to disruption of the SMN locus,^5^ most commonly due to deletions or gene conversions resulting in *SMN1* deficiency. The SMN locus consists of two nearly identical paralogs, *SMN1* and *SMN2*. The ‘C’ nucleotide at the c.840 position of *SMN1* facilitates proper splicing, while the ‘T’ at that position in *SMN2* causes exon 7 skipping in 85% of *SMN2* transcripts (**Figure 1A**). As a result, *SMN2* alone does not produce a sufficient amount of functional SMN protein, causing neurons that have zero functional copies of *SMN1* to die prematurely. In the general population, individuals typically inherit two intact copies of each paralog, though genomes with between zero and five copies of each have been observed.^3^ Since *SMN2* can still produce a small amount of SMN protein, disease severity and age of onset in affected individuals is influenced by the copy number of *SMN2*.

**Figure 1.**
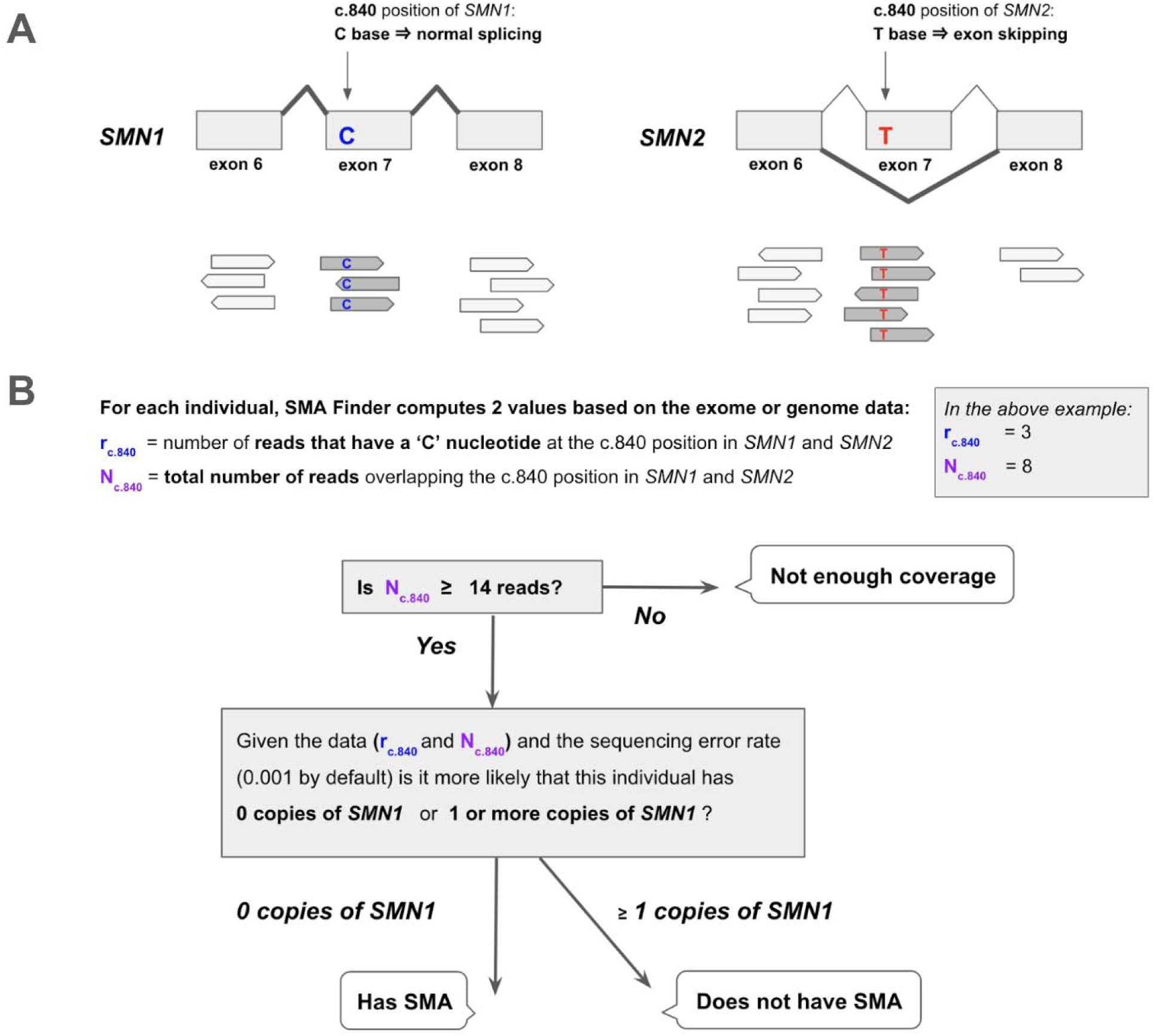
Detecting SMA using reads aligned to the *SMN1* and *SMN2* paralogs. **A.** The *SMN1* and *SMN2* paralogs are 99.9% identical. One of the few differences between them occurs at their c.840 position. The ‘C’ at this position in *SMN1* leads to proper splicing, while the ‘T’ in *SMN2* leads to skipping of exon 7 in most *SMN2* transcripts. Individuals that have zero functional copies of *SMN1* develop spinal muscular atrophy (SMA), and the severity of their disease is inversely proportional to the number of copies of *SMN2* in their genome since each copy of *SMN2* can produce a small amount of SMN protein. **B.** SMA Finder works by counting all aligned reads that overlap the c.840 position in both *SMN1* and *SMN2* and then computing the fraction of reads that have a ‘C’ at that position. This fraction is interpreted as the fraction of intact *SMN1* copies in the individual’s genome. When it is near zero, it implies the absence of any functional copies of *SMN1*, and therefore suggests that the sample is positive and the individual has a diagnosis of SMA.

The near-perfect sequence homology between *SMN1* and *SMN2* causes read alignment algorithms like BWA^6^ to either mismap or ambiguously map reads at this locus, often resulting in reads having a mapping quality of 0. This, in turn, confounds standard variant calling pipelines such as GATK^7^, and prevents them from being directly useful for SMA diagnosis. To address these issues, multiple specialized tools have been developed to diagnose SMA from genome and long read sequencing data, including SMNCopyNumberCaller^3^ and Paraphase.^8^ Additionally, the MYO-SEQ study^9^ published an SMA calling pipeline for exome data on GRCh37 and used it to identify 5 novel SMA diagnoses in a cohort of 1001 cases with limb-girdle weakness. Most recently, the Chameleolyser tool was described in a study involving GRCh37-aligned exome samples from a cohort of 17,650 undiagnosed patients.^10^ There, Chameleolyser successfully identified 15 novel SMA cases. However, to our knowledge, no publicly available tool exists for detecting SMA in exome or panel sequencing samples on GRCh38 or the new telomere-to-telomere (T2T-CHM13)^11^ reference. We therefore created SMA Finder for use with exome, genome, or panel sequencing data aligned to GRCh37, GRCh38, or T2T-CHM13, and applied it to diagnose unsolved cases within large rare disease cohorts from the Broad Institute Center for Mendelian Genomics (CMG) and Tartu University Hospital.

## Materials and Methods

### SMA Finder algorithm

SMA Finder works by computing two numbers from the read data, **r** and **N**, where **N** is the total number of aligned reads that overlap the c.840 position of *SMN1* and *SMN2*, and **r** is the subset of these reads that have a ‘C’ at the c.840 position. When there is sufficient read coverage (**N** ≥ 14), SMA Finder interprets the lack of reads with a ‘C’ at c.840 as evidence that the individual has zero functional copies of *SMN1* (**Figure 1B**, **Supplementary Methods**), and so reports a positive call. **Figure 3** shows how **r** and **N** are used to distinguish between true positive and true negative samples.

### Cohort summary

The Broad Institute Center for Mendelian Genomics (CMG) cohort consisted of 20,537 individuals from 10,754 families with a wide spectrum of medical conditions. Families sequenced through the CMG were enrolled at the Broad Institute by the Rare Genomes Project or enrolled in research studies with local regulatory approval through the participating collaborators, including for sharing de-identified samples for sequencing and analysis. This project was approved by the Mass General Brigham IRB (protocols #2016P001422 and #2013P001477). In this cohort, 12,045 (59%) individuals were affected with a variety of suspected monogenic disorders, while 8,401 (41%) were unaffected parents or relatives (**Figure 2A**). We performed ancestry inference for 20,205 individuals by computing principal components for high-quality bi-allelic autosomal SNVs using the gnomAD v2 method.^12^ We found that 13,240 (66%) individuals were European and the remaining 34% came from various other populations (**Figure 2B**). Among the 12,045 affected individuals, 10,125 (84%) had phenotype descriptions encoded in Human Phenotype Ontology (HPO) terms.^13^ Of these, 5,138 (51%) had at least one HPO term in the “Abnormality of the nervous system” category, and 4,035 (40%) had at least one HPO term in the “Abnormality of the musculoskeletal system” category (**Figure 2C**).

**Figure 2.**
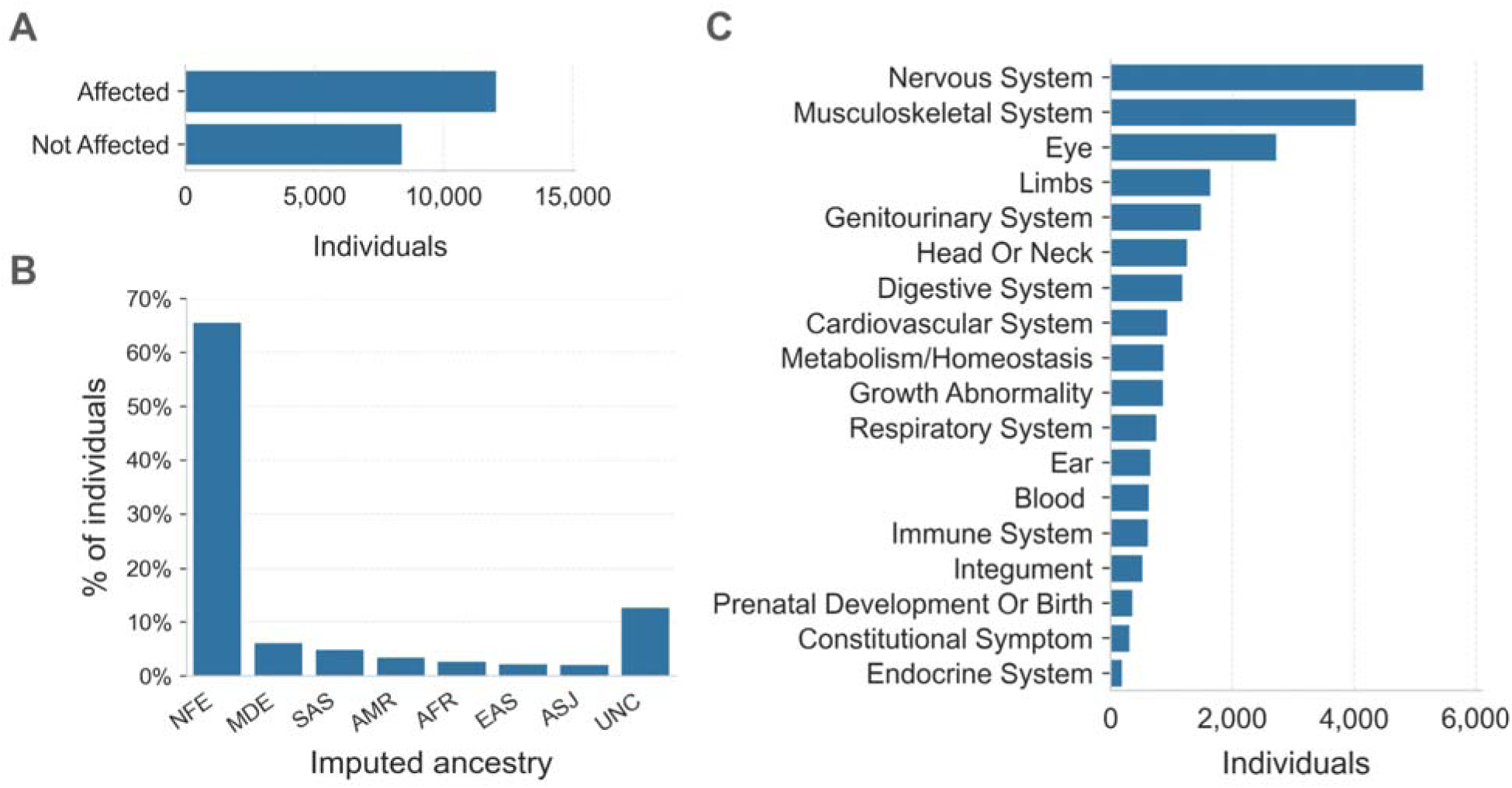
Overview of the CMG rare disease cohort. **A.** The affected status of individuals in the CMG cohort is shown on the y-axis. 12,045 individuals are in the Affected category, 8,401 are Not Affected, and 91 individuals have unknown affected status. Here “Affected” means that the individual was enrolled in a rare disease cohort due to having a disease considered to be rare and most likely genetic in origin. **B.** Inferred ancestry of individuals within the CMG cohort is shown on the x-axis: NFE (Non-Finnish Europeans), MDE (Middle Eastern), SAS (South Asian), AMR (Admixed American), AFR (African/African American), EAS (East Asian), ASJ (Ashkenazi Jewish), and UNC (unclassified). **C.** The top-level categories from the Human Phenotype Ontology (HPO) are shown on the y-axis. Any individual with multiple HPO terms was counted only once in each category but may be counted more than once across categories.

**Fig 3.**
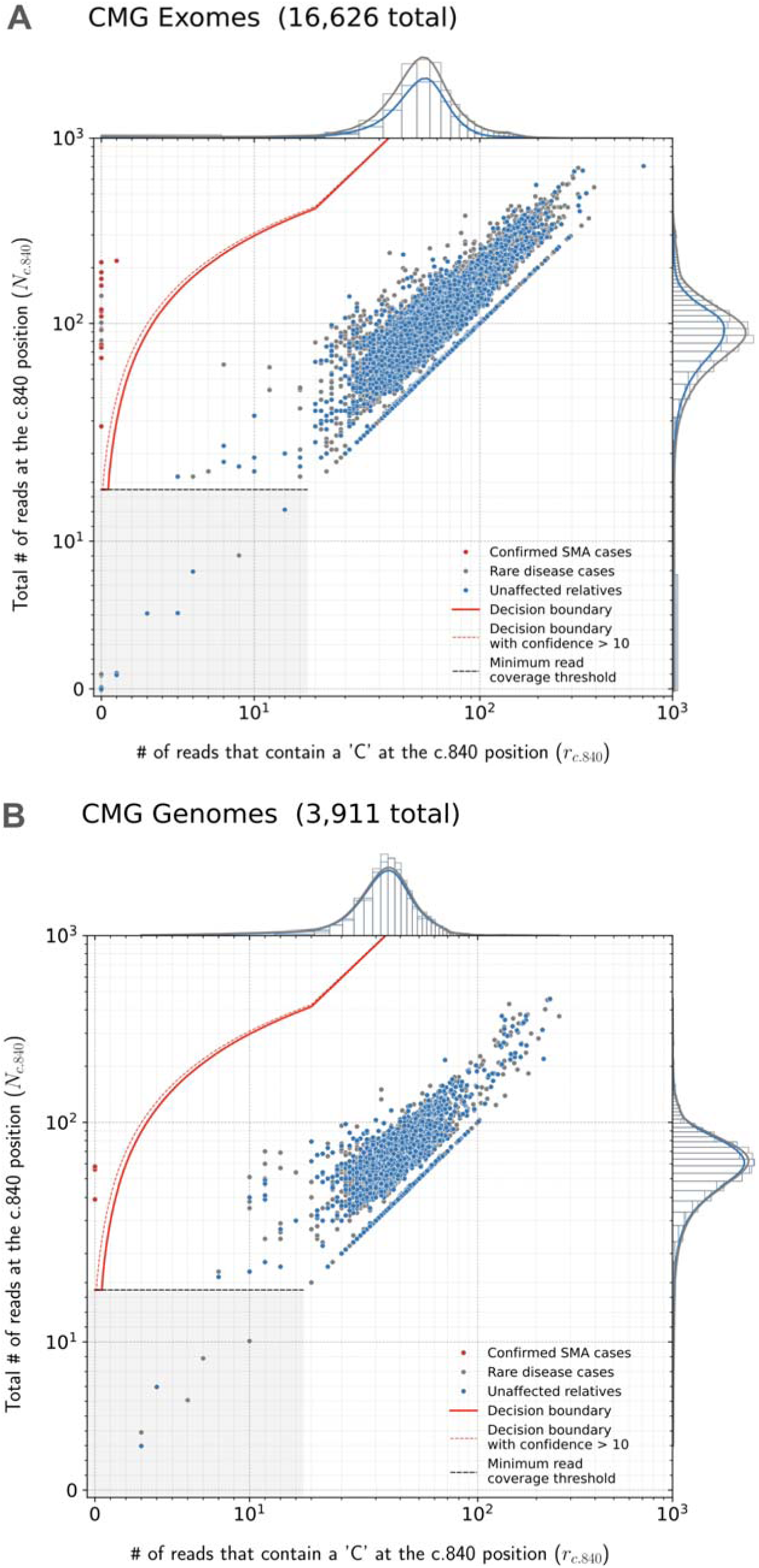
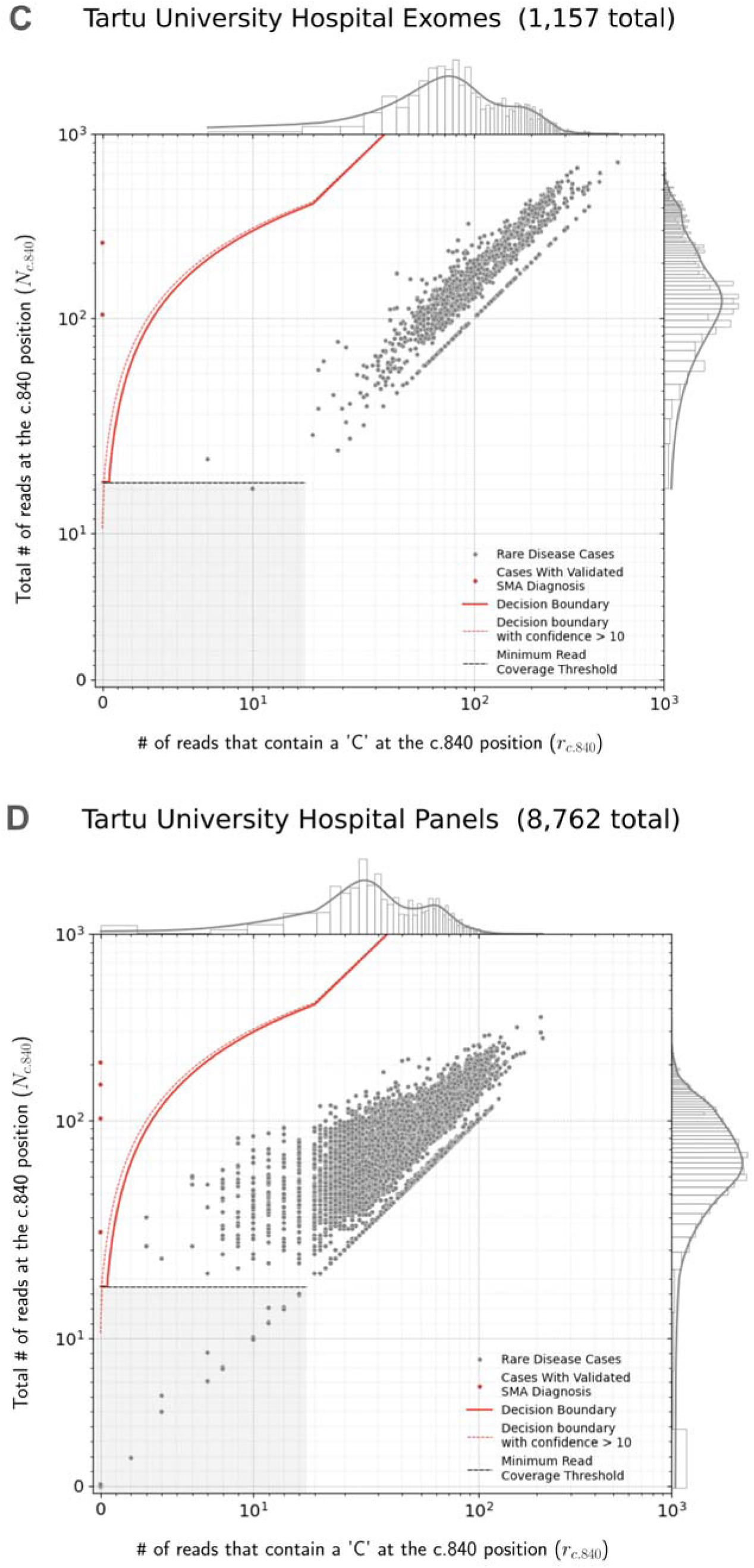
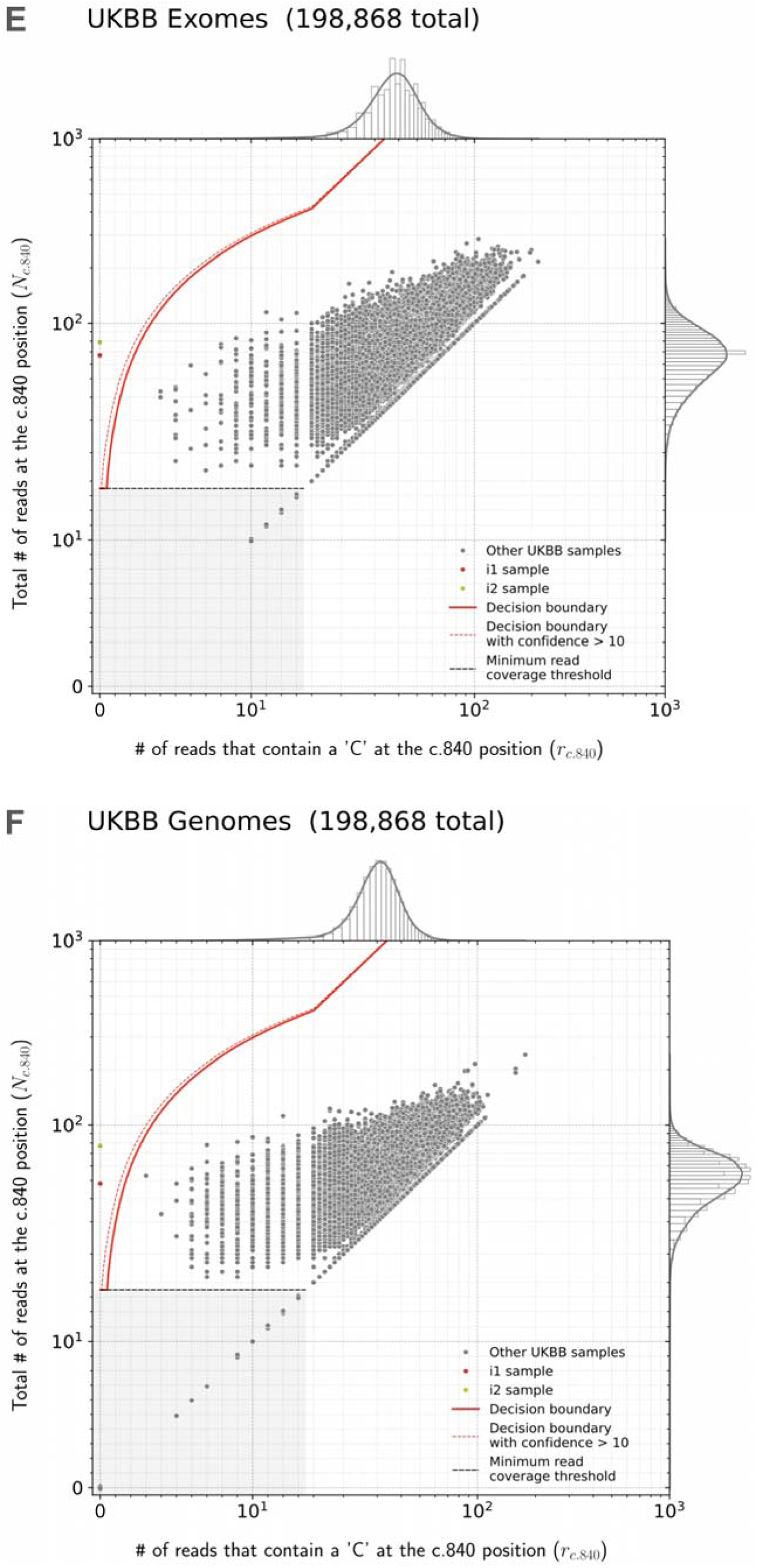
SMA Finder results. Read counts measured by SMA Finder in exome (**A**) and genome (**B**) samples from CMG cohorts, as well exomes (**C**) and panel sequencing samples (**D**) from Tartu University Hospital. Each dot represents a sample. The red line represents the decision boundary used by SMA Finder which reports samples to the left of the boundary as SMA-positive. Samples in the gray box where y ≤ 14 are reported as having insufficient read coverage to make a call. The red dots represent previously known SMA diagnoses, the gray dots are rare disease cases (including the new SMA diagnoses), and the blue dots are unaffected relatives. To clearly show points across a large range of read count values, the x and y axes use a symmetrical log scale that is linear in the range 0 ≤ x ≤ 14 and 0 ≤ y ≤ 14 before switching to a logarithmic scale for x or y > 14. This choice of scale causes part of the decision boundary to appear curved even though it is linear in standard Cartesian coordinates. **E** and **F** show SMA Finder read counts for 198,868 UKBB exomes and genomes respectively. The red dot represents UKBB sample i1 which had phenotype records consistent with an SMA diagnosis and was called positive by both SMA Finder and SMNCopyNumberCaller. The yellow dot represents i2 which was only called positive by SMA Finder and was a no-call from SMNCopyNumberCaller. Marginal histograms show the density of scatter plot points along each axis, with the histogram along the vertical axis showing a distribution of read counts overlapping the c.840 position in *SMN1* + *SMN2*, while the histogram along the horizontal axis shows the number of reads with a ‘C’ at the c.840 position. **NOTE:** The exome, genome, and panel sequencing samples in **A** and **B** as well as in **C** and **D** are largely from non-overlapping sets of individuals, while the exomes and genomes in **E** and **F** are alternative samples from the same set of 198,868 individuals in UKBB.

Genome sequencing used PCR-free library preparation and Illumina HiSeq X Ten v2 chemistry to generate 150bp paired-end reads. The mean target coverage was >30x. Exome sequencing used Illumina Nextera or Twist exome capture (∼38 Mb target) and similarly generated 150bp paired-end reads. It aimed to cover >80% of targets at 20x and a mean target coverage of >60x. Both genome and exome data were processed using GATK best practices, starting with BWA alignment to GRCh38 and GATK base quality score recalibration (BQSR). We used Hail Batch^14^ to run SMA Finder on all samples in parallel. The average number of reads overlapping the c.840 position of *SMN1* + *SMN2* was 77.9 across CMG genomes and 110.7 across CMG exomes. These numbers reflect the higher coverage in exomes compared to genomes at this locus, as well as the total copy number of SMN paralogs per individual.

The Tartu University Hospital cohort consisted of 9,919 pseudonymised samples sent for molecular diagnostics for various suspected medical conditions to Tartu University Hospital between 2015 - 2022. The cohort analysis was approved by the University of Tartu Research Ethics Committee (protocol # 374M-6). Out of the 9,919 samples, 1,157 (11.7%) were exomes and 8,762 (88.3%) were panel sequencing panels generated using the Illumina TruSight™ One or the Illumina TruSight™ One Expanded panel of 4,811 to 6,794 genes. Guaranteed mean coverage for both panels and exomes was >100x for panel regions. The data was aligned to reference genome build GRCh37 using BWA. Samples were annotated with summary clinical characteristics, and de-pseudonymized data was accessed for positive SMA Finder results only.

The UK Biobank (UKBB)^15^ dataset included de-identified genetic and medical records from 198,868 individuals as well as both exome and genome samples for each individual. These samples were aligned to GRCh38 using BWA. The UKBB is a large population-based prospective study that recruited UK residents between 40 and 69 years of age during the years 2006–2010. UKBB genotypes had a mean depth of coverage of 32.5x and a minimum of 23.5x.^16^ For UKBB exomes, the depth of coverage, on average, exceeded 20x at 95% of panel bases.^17^ The average number of reads overlapping the c.840 position of *SMN1* + *SMN2* was 56.8 in UKBB genomes and 75.1 in UKBB exomes, again reflecting the higher coverage in exomes at this locus, as well as the total copy number of SMN paralogs per individual.

Our cohorts did not include any samples aligned to the T2T-CHM13 reference. Therefore, to test SMA Finder’s performance on T2T-CHM13, we realigned the 21 confirmed-positive exome samples from the CMG cohort to the T2T-CHM13 reference using BWA MEM and ran SMA Finder on the realigned samples. We then repeated these steps on 21 randomly selected exomes from the CMG cohort that SMA Finder had previously called as SMA-negative. After realignment to T2T-CHM13, SMA Finder still reported the 21 positive samples as SMA-positive, and the 21 negative samples as SMA-negative, confirming its utility for T2T-aligned samples.

## Results

We applied SMA Finder to 16,626 exomes and 3,911 genomes from phenotypically heterogeneous rare disease cohorts within the Broad CMG,^18^ part of the GREGoR Consortium. SMA Finder identified all 13 known SMA-positive samples (10 exomes and 3 genomes), and flagged 10 previously undiagnosed exome samples as candidate SMA cases, of which 8 have now been validated by gold standard methods such as MLPA. For the remaining novel diagnoses (n=2), confirmatory testing is pending in one case, and is not possible in the other due to loss of contact with the patient (**Table 1**). SMA Finder was negative for 8,401 unaffected samples (6,500 exomes and 1,901 genomes). SMA Finder reported insufficient read coverage to make a call in 112 exomes (0.7%) and 6 genomes (0.2%) due to these samples having fewer than 14 total reads overlapping c.840 in *SMN1* + *SMN2*. Strikingly, out of the 23 total SMA-positive cases in this heterogeneous rare disease cohort, 20 had an initial clinical diagnosis of muscular dystrophy or myopathy while one had a clinical diagnosis of congenital myasthenic syndrome.

**Table 1:** Positive cases identified by SMA Finder in the rare disease cohorts. Table 1 lists all samples called as SMA-positive by SMA Finder within the Broad Institute Center for Mendelian Genomics (CMG) and Tartu University Hospital cohorts. The Original Clinical Diagnosis column lists the clinical diagnosis prior to SMA testing. The SMA Confirmation Status column shows whether the diagnosis was already known at the time of the SMA Finder result as well as the status of confirmatory testing for novel diagnoses. Age of Onset and SMA Type columns are based on physician reports, while the SMN2 Copies column is based on the SMN2 copy number as measured by MLPA genetic testing. The next three columns display the confidence score, **r**, and **N** counts computed by SMA Finder (see Figure1, Methods), while the last column shows the imputed ancestry as NFE (Non-Finnish European), MDE (Middle Eastern), AFR (African/African American), SAS (South Asian), or UNC (unclassified).

|  | Sample Id | Cohort | Type | Original Clinical Diagnosis | SMA Confirmation Status | Age of Onset | SMA Type | SMN2 Copies | Confidence Score | Reads With 'C' at c.840 | Total Reads at c.840 | Imputed Ancestry |
| --- | --- | --- | --- | --- | --- | --- | --- | --- | --- | --- | --- | --- |
| G1 | RGP_1153_3 | CMG | genome | limb-girdle muscular dystrophy | Previously known confirmed diagnosis: positive control |  |  |  | 56 | 0 | 58 | NFE |
| G2 | RGP_823_3 | CMG | genome | muscular dystrophy | Previously known confirmed diagnosis: positive control |  |  |  | 37 | 0 | 39 | NFE |
| G3 | RGP_916_3 | CMG | genome | spinal muscular atrophy | Previously known confirmed diagnosis: positive control |  |  |  | 54 | 0 | 56 | NFE |
| E1 | MAAC057 | CMG | exome | limb-girdle muscular dystrophy | Previously known confirmed diagnosis: positive control |  |  |  | 74 | 0 | 77 | NFE |
| E2 | MANT033 | CMG | exome | limb-girdle muscular dystrophy | Previously known confirmed diagnosis: positive control |  |  |  | 207 | 0 | 214 | NFE |
| E3 | MCAI001 | CMG | exome | limb-girdle muscular dystrophy | Previously known confirmed diagnosis: positive control |  |  |  | 168 | 0 | 174 | MDE |
| E4 | MNOT001 | CMG | exome | limb-girdle muscular dystrophy | Previously known confirmed diagnosis: positive control |  |  |  | 182 | 0 | 189 | NFE |
| E5 | MPAU019 | CMG | exome | limb-girdle muscular dystrophy | Previously known confirmed diagnosis: positive control |  |  |  | 90 | 0 | 94 | NFE |
| E6 | MLEU047 | CMG | exome | limb-girdle muscular dystrophy | Previously known confirmed diagnosis: positive control |  |  |  | 154 | 0 | 160 | UNC |
| E7 | SCO_PED004D_GA0014_D1 | CMG | exome | neonatal death | Previously known confirmed diagnosis: positive control |  |  |  | 27 | 0 | 28 | UNC |
| E8 | MLEU044 | CMG | exome | limb-girdle muscular dystrophy | Previously known confirmed diagnosis: positive control |  |  |  | 105 | 0 | 109 | NFE |
| E9 | MPAU042 | CMG | exome | limb-girdle muscle weakness | Previously known confirmed diagnosis: positive control |  |  |  | 71 | 0 | 74 | UNC |
| E10 | MGLA019 | CMG | exome | limb-girdle muscular dystrophy | Previously known confirmed diagnosis: positive control |  |  |  | 97 | 0 | 101 | NFE |
| E11 | RD6237 | Tartu | exome | spinal muscular atrophy I, epilepsy | Previously known confirmed diagnosis: positive control |  |  |  | 247 | 0 | 256 | NFE |
| T1 | IT334 | Tartu | panel | spinal muscular atrophy I | Previously known confirmed diagnosis: positive control |  |  |  | 24 | 0 | 25 | NFE |
| T2 | NV919 | Tartu | panel | limb-girdle muscular dystrophy | Previously known confirmed diagnosis: positive control |  |  |  | 197 | 0 | 204 | NFE |
| T3 | VH4169 | Tartu | panel | myopathy | New diagnosis: confirmed via MLPA | 3 | III | 4 | 98 | 0 | 102 | NFE |
| T4 | KT6879 | Tartu | panel | moderate intellectual disability | New diagnosis: confirmed via MLPA. Incidental finding. | asymptomatic at 14 | IV? | 5 | 149 | 0 | 155 | NFE |
| E12 | YE8324 | Tartu | exome | mild proximal muscle weakness | New diagnosis: confirmed via MLPA | <10 | III | 4 | 100 | 0 | 104 | NFE |
| E13 | MBEL164 | CMG | exome | limb-girdle muscular dystrophy | New diagnosis: confirmed via MLPA | 13 | III | 4 | 113 | 0 | 117 | NFE |
| E14 | BON_B22_41_1_D1 | CMG | exome | muscular dystrophy | New diagnosis: confirmed via MLPA | <14 | III | 4 | 115 | 0 | 119 | NFE |
| E15 | MBRN009 | CMG | exome | limb-girdle muscular dystrophy | New diagnosis: confirmed via MLPA | 3 | III | 4 | 89 | 0 | 93 | NFE |
| E16 | UWA_LAI8985_LAI013-1363_1 | CMG | exome | congenital myopathy | New diagnosis: confirmed via MLPA | 0 | I | 2 | 62 | 0 | 65 | UNC |
| E17 | MTAM002 | CMG | exome | limb-girdle muscle weakness | New diagnosis: confirmed via MLPA | 2 | III | 4 | 112 | 0 | 116 | MDE |
| E18 | MTAM032 | CMG | exome | limb-girdle muscular dystrophy | New diagnosis: confirmed via PCR showing homozygous loss of SMN1 exon 7 | 4 | III |  | 78 | 0 | 81 | UNC |
| E19 | MWAR098 | CMG | exome | limb-girdle muscular dystrophy | New diagnosis: confirmed via MLPA | 1 | III | 3 | 182 | 1 | 218 | NFE |
| E20 | D304-13N-TB220379 | CMG | exome | limb-girdle muscular dystrophy | New diagnosis: confirmed via MLPA | mid teens | III | 4 | 217 | 0 | 225 | NFE |
| E22 | TOP_N799366_1_1 | CMG | exome | congenital myasthenic syndrome | New diagnosis: clinical correlation confirmed, genetic confirmation in progress | 6 | III |  | 97 | 0 | 101 | SAS |
| E21 | K0004_01_1 | CMG | exome | limb-girdle muscular dystrophy | New diagnosis: clinical correlation confirmed, genetic confirmation not possible | 13 | III |  | 136 | 0 | 141 | AFR |
|  |  |  |  |  |  |  |  |  | 24 | 0 | 25 = Min |  |
|  |  |  |  |  |  |  |  |  | 100 | 0 | 104 = Median |  |
|  |  |  |  |  |  |  |  |  | 247 | 1 | 256 = Max |  |

To evaluate its performance beyond the CMG dataset, we also ran SMA Finder on 9,919 samples (1,157 exomes and 8,762 panel sequencing samples) from another heterogeneous rare disease cohort at the Tartu University Hospital. SMA Finder detected three previously known cases and three new cases which have since been confirmed by clinical testing (**Table 1**). Of the three new cases, two presented as SMA type III, and one had symptoms that did not include obvious SMA features and thus may be a presymptomatic case of SMA type IV.

Finally, to measure SMA Finder’s positive predictive value (PPV) in a large population cohort as well as compare its performance to other tools and across sample types, we applied SMA Finder to 198,868 individuals from the UK Biobank (UKBB).^15^ Given the availability of an exome and genome sample for each individual, we ran SMA Finder on all exomes and genomes, while also running SMNCopyNumberCaller^3^ on the genomes. We found SMA Finder calls to be identical between exomes and genomes, with the only difference being that SMA Finder reported 85 out of 198,868 (0.042%) genome samples and 14 out of 198,868 (0.007%) exome samples (Fisher’s exact p=1.4x10^-13^) as having insufficient read coverage to make a call (**Table S3**). SMA Finder flagged two individuals (i1 and i2) as SMA-positive, with their exome and genome samples yielding the same result. It reported all other exomes and genomes that had sufficient coverage as SMA-negative (**Figure 3E, F**). Concordance between SMA Finder and SMNCopyNumberCaller was also nearly 100%. Like SMA Finder, SMNCopyNumberCaller (a genome-only tool) reported individual i1 to be SMA-positive, while reporting no-call for 35 genomes including i2 (**Table S2**).

The UKBB phenotype records for i1 included ICD-10 code G121 “Other inherited spinal muscular atrophy”, suggesting this to be a true positive call for both SMA Finder and SMNCopyNumberCaller. Additionally, i1’s records indicated that they died between the age of 65 and 70, with the cause of death listed as “Spinal muscular atrophy”. In contrast, i2 had few phenotype records, making it difficult to tell whether this was a false positive call by SMA Finder, or a missed call by SMNCopyNumberCaller. The fact that SMA Finder made the same call for both the exome and genome of i2 increases the likelihood that this is a true positive. However, even if we conservatively count i2 as a potential false positive call by SMA Finder based on the absence of phenotype records and the no-call result from SMNCopyNumberCaller, SMA Finder’s overall false positive rate would still be < 1 / 200,000 exomes. Additionally, this would yield a PPV of 97% based on 28 true positive samples (21 from CMG cohorts, 6 from Tartu University Hospital, and 1 from UKBB) and at most 1 hypothetical false positive (counting sample i2 from UKBB).

## Discussion

We developed and validated SMA Finder, a new tool for detecting SMA-positive samples within short read exome, genome, and panel sequencing data. After testing this tool on multiple heterogeneous rare disease cohorts, as well as nearly 200,000 individuals from the UKBB, we found its false positive rate to be less than 1 in 200,000. Our results showed that SMA Finder is a robust and accurate tool for detecting the most common molecular causes of SMA using exome, genome, and panel sequencing samples.

However, the analysis had several limitations related to these cohorts. It was predominantly based on samples of European ancestry. A precise estimate of SMA Finder’s accuracy in other populations will require testing on cohorts with larger numbers of non-European samples. Similarly, since most samples used DNA extracted from blood, further testing is needed to measure SMA Finder’s accuracy for other sample sources such as saliva or dried blood spots.

The SMA Finder algorithm in its current form also has several important limitations. First, it only determines whether an individual has zero or more than zero functional copies of *SMN1*, and thus does not provide information on carrier status or *SMN2* copy number. In the future, we may extend SMA Finder to estimate the exact copy numbers of *SMN1* and *SMN2*. Second, SMA Finder only tests for variants that disrupt the c.840 position of *SMN1*. Studies have shown that, depending on ancestry, between 6% and 49% of SMA cases^19^ may be caused by other variants in regions of the *SMN1* gene that don’t overlap the c.840 position. SMA Finder cannot currently detect these variants, and so would yield false negative results in those cases.

Furthermore, SMA Finder was unable to make a call in 112 exomes (0.7%) and 6 genomes (0.2%) from the CMG as well as 14 exomes (0.007%) and 85 genomes (0.04%) from UKBB due to insufficient read coverage at the c.840 position. These no-call percentages may be useful for confirming sequencing data quality when applying SMA Finder to new cohorts where a significantly higher proportion of no-calls would indicate problems with target capture or sequencing at the SMN locus. Finally, SMA Finder was designed and tested on samples generated using Illumina sequencers and aligned using BWA.^6^ Samples generated using a different sequencing technology, or aligned using a different, non-functionally-equivalent aligner may not work as expected, particularly when aligning to the GRCh38 reference where the ALT contigs include additional copies of *SMN1* and *SMN2* (**Supplementary Methods**).

Despite these limitations, SMA Finder’s exceptionally high accuracy in detecting homozygous loss of SMN1 due to deletions or gene conversions at the c.840 position allowed us to shed new light on open questions about the genetic architecture of SMA. Over the years, reports have described cases of unaffected individuals with biallelic loss of SMN1 due to deletions that would be detectable by SMA Finder.^20,21,22^ However, the frequency of such cases in the general population has not been well-characterized. Our analysis of UKBB exomes and genomes using SMA Finder and SMNCopyNumberCaller shows that such cases are extremely rare in the European population, occurring at a frequency below 1 in 200,000.

Rare disease diagnostic pipelines based on short read sequencing data have, until recently, needed to exclude highly repetitive regions such as segmental duplications due to technical limitations of existing variant calling algorithms. However, our analysis adds to the growing body of research ^3,10,23^ which demonstrates that, with focused tool development, it is possible to detect variants in these regions with sufficient accuracy for rare disease diagnosis. Furthermore, at some loci such as the SMN c.840 position, our ability to accurately detect the causal variant from exome or panel sequencing data can be comparable to the accuracy of variant calling in non-repetitive regions.

A striking observation from our cohorts was that 11 out of the 13 new SMA diagnoses were made in individuals carrying a clinical diagnosis of muscular dystrophy, congenital myasthenia, or myopathy, and 12 of the 16 known SMA cases carried an initial clinical misdiagnosis of muscular dystrophy, reinforcing similar observations by other groups.^24^ Although it is known that the differential diagnosis of an infant or adolescent with progressive weakness includes SMA in addition to muscular dystrophies, myopathies, and congenital myasthenic syndromes^25^, clinical gene panels for muscular dystrophy and myopathy may not include SMA testing. Moreover, clinicians and researchers may be misled by the presence of variants of unknown significance (VUS) in other neuromuscular disease genes, by complementary diagnostic tools such as electromyography (EMG) and muscle MRI, and by the highly elevated creatine kinase (CK) levels sometimes observed in ambulant patients with SMA type III. Prior to the inclusion of SMA testing in newborn screening, there was commonly a diagnostic delay in SMA, particularly for the milder types II and III which can present with clinical features that are similar to later-onset Mendelian myopathies.^26^ Now, with newly-available SMA treatments where early detection is imperative for the best therapeutic effect,^29^ there is increased urgency to address diagnostic delays. Our analysis demonstrates that the inclusion of an SMA detection tool within existing rare disease analysis pipelines - particularly for cohorts that include neuromuscular phenotypes - can successfully identify missed cases.

In 2021, the American College of Medical Genetics (ACMG) published an updated set of recommendations for reporting of secondary findings (SF) in clinical exome and genome sequencing.^28^ In the section titled "Poor candidates for secondary findings, due to concerns about analytical validity", the authors listed reasons why genes with homologous sequences should be excluded from the SF list. These included concerns about the accuracy of variant detection in these regions as well as challenges in orthogonal validation. Given the clinically overlooked diagnoses presented in this paper, the demonstrated PPV of SMA Finder, as well as the widespread availability of confirmatory molecular testing for SMA, we propose that *SMN1* should now be considered for inclusion in the ACMG SF list.

### Declaration of interests

HLR receives research funding from Microsoft and previously received funding from Illumina to support rare disease gene discovery and diagnosis. AODL has consulted for Tome Biosciences, Ono Pharma USA Inc, and Addition Therapeutics, and is member of the scientific advisory board for Congenica Inc and the Simons Foundation SPARK for Autism study. AL received honoraria for speaking at educational events for Biogen, PTC and Roche, is a subinvestigator in clinical trials by Roche and PTC, and is involved in a project supported by Biogen (POL-SMA-17-11166). PBK has received research support from ML Bio and Sarepta Therapeutics, and has consulted for Lupin, Neurogene, NS Pharma, and Teneofour.

## Data Availability

Genomic and phenotypic data for GREGoR and CMG cohorts is available via dbGaP accession numbers phs003047 and phs001272. Access is managed by a data access committee designated by dbGaP and is based on intended use of the requester and allowed use of the data submitter as defined by consent codes. The Tartu University cohort is not publicly available. UKBB data is accessible to researchers who obtain Tier 3 access to the UK Biobank.

## Acknowledgments

This paper is dedicated to Clara and Dmitry Bogomolny. We thank the many families who participate in these rare disease studies and the Broad CMG collaborators for sharing the samples and medical data. The Broad CMG sequencing and analysis was funded by the National Human Genome Research Institute (NHGRI), the National Eye Institute, the National Heart, Lung and Blood Institute grant UM1HG008900 and NHGRI grants U01HG011755 and R01HG009141. GH is supported by the GREGoR Consortium, and research in this publication was supported by the National Human Genome Research Institute of the National Institutes of Health under Award Number U24HG011746. This publication has also been made possible in part by CZI grants 2019-19927, 2020-224274, and 2022-309464 https://doi.org/10.37921/236582yuakxy, from the Chan Zuckerberg Initiative DAF, an advised fund of Silicon Valley Community Foundation (funder DOI 10.13039/100014989). The Tartu University Hospital cohort analysis was funded by the Estonian Research Council grants PSG774 and PRG471. VSG was supported by NIH NHGRI grant T32HG010464. Work in CGB’s laboratory is supported by intramural funds of NINDS/NIH. This research has been conducted using the UK Biobank Resource under application number 48511. The content is solely the responsibility of the authors and does not necessarily represent the official views of the National Institutes of Health or the other organizations that have funded the work. HL receives support from the Canadian Institutes of Health Research (CIHR) for Foundation Grant FDN-167281 (Precision Health for Neuromuscular Diseases), Transnational Team Grant ERT-174211 (ProDGNE) and Network Grant OR2-189333 (NMD4C), from the Canada Foundation for Innovation (CFI-JELF 38412), the Canada Research Chairs program (Canada Research Chair in Neuromuscular Genomics and Health, 950-232279), the European Commission (Grant # 101080249) and the Canada Research Coordinating Committee New Frontiers in Research Fund (NFRFG-2022-00033) for SIMPATHIC, and from the Government of Canada Canada First Research Excellence Fund (CFREF) for the Brain-Heart Interconnectome (CFREF-2022-00007). KP is a recipient of CIHR Postdoctoral fellowship (202210MFE-491707-404816). RH is supported by the Wellcome Discovery Award (226653/Z/22/Z), the Medical Research Council (UK) (MR/V009346/1), Ataxia UK, Action for AT, the Muscular Dystrophy UK, the LifeArc Centre to Treat Mitochondrial Diseases (LAC-TreatMito), the UKRI/Horizon Europe MSCA Doctoral Network Programme (Project 101120256 — MMM) and an MRC strategic award to establish an International Centre for Genomic Medicine in Neuromuscular Diseases (ICGNMD) MR/S005021/1. This research was supported by the NIHR Cambridge Biomedical Research Centre (BRC-1215-20014). The views expressed are those of the authors and not necessarily those of the NIHR or the Department of Health and Social Care.

## Data and code availability

SMA Finder is publicly available as a stand-alone tool at https://github.com/broadinstitute/sma_finder under the open-source MIT license.

Also, it is available as a WDL workflow on Terra at https://portal.firecloud.org/?return=terra#methods/translational-genomics-group/sma-finder/4

## Supplementary Materials

### Installing and Running SMA Finder

The SMA Finder tool is implemented in python v3.7+ and can be installed by running

`python3 -m pip install sma_finder`

The --help option can be used to print out SMA Finder’s command line arguments:

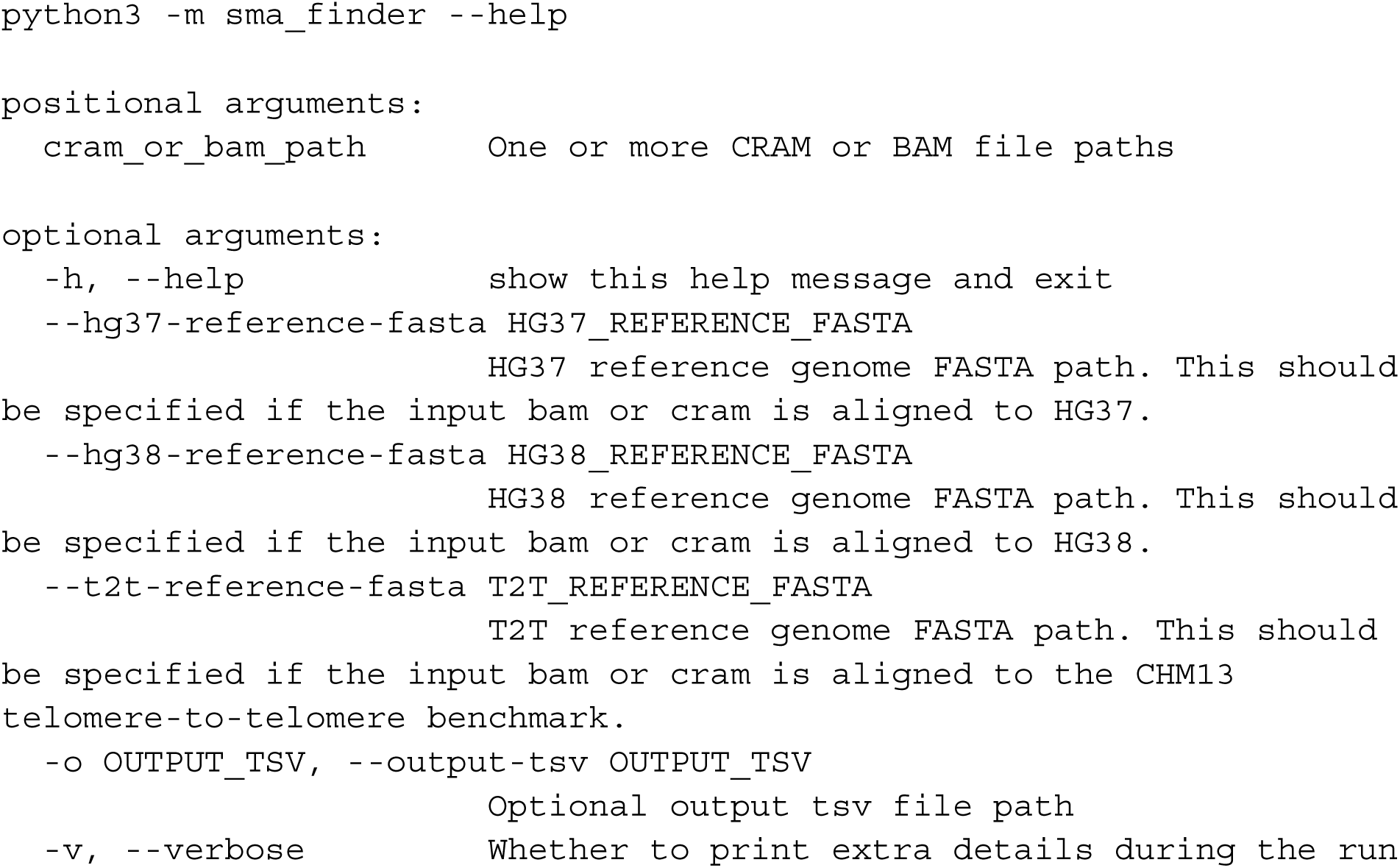

SMA Finder has two required inputs:

1) a reference genome file provided via the appropriate `--*-reference-fasta` option
2) the path(s) of one or more aligned read data files in bam or cram format

It outputs a tab-separated table with one row per input file with the following columns:

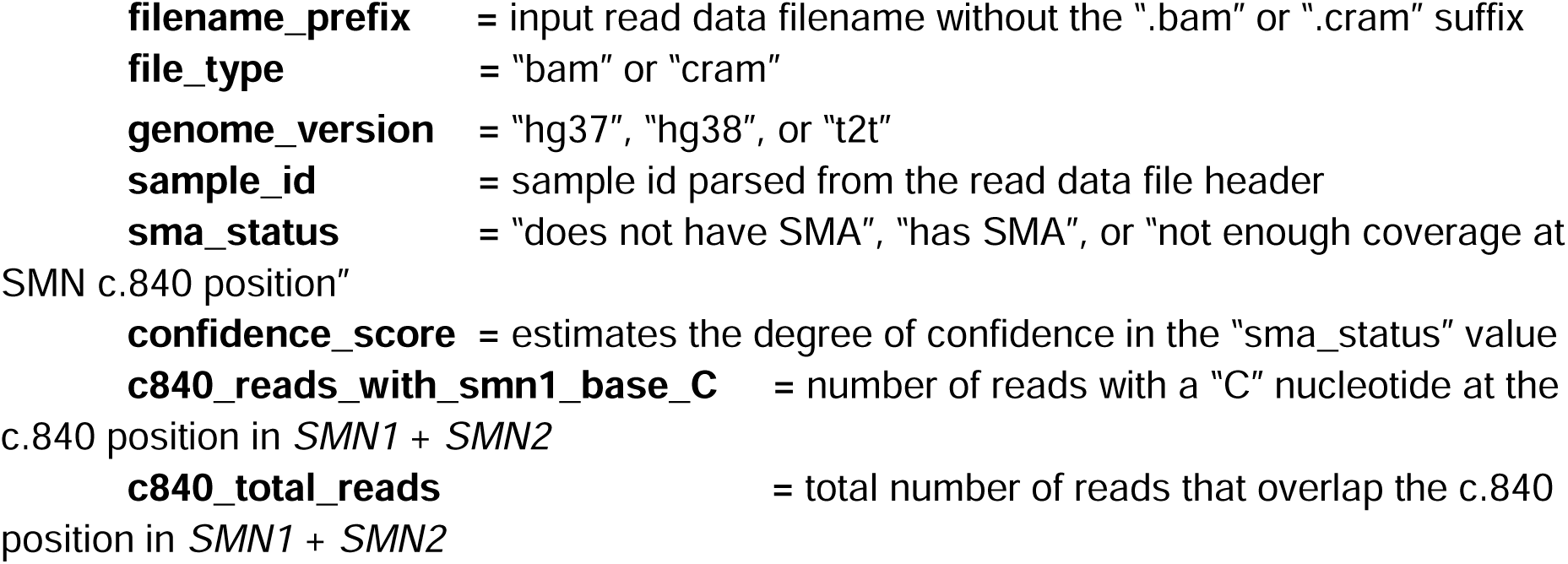

**Table S1.** Example SMA Finder output table.

| filename_prefix | file_type | genome_version | sample_id | sma_status | confidence_score | c840_reads_with_smn1_base_C | c840_total_reac |
| --- | --- | --- | --- | --- | --- | --- | --- |
| sample1 | bam | hg38 | s5 | does not have SMA | 2524 | 85 | 146 |
| sample2 | cram | hg38 | MWAR098 | has SMA | 182 | 1 | 218 |
| sample3 | cram | hg37 | s12421 | not enough coverage at SMN c.840 position | 0 | 2 | 8 |

**Table S2.**
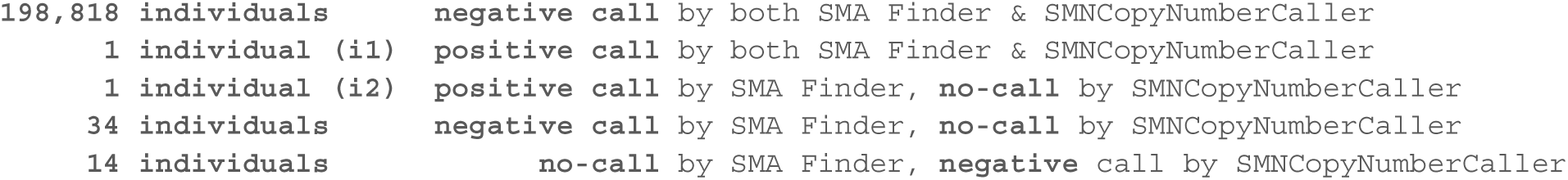
Concordance between SMA Finder exome calls vs SMNCopyNumberCaller genome calls in 198,868 UKBB participants.

**Table S3.**
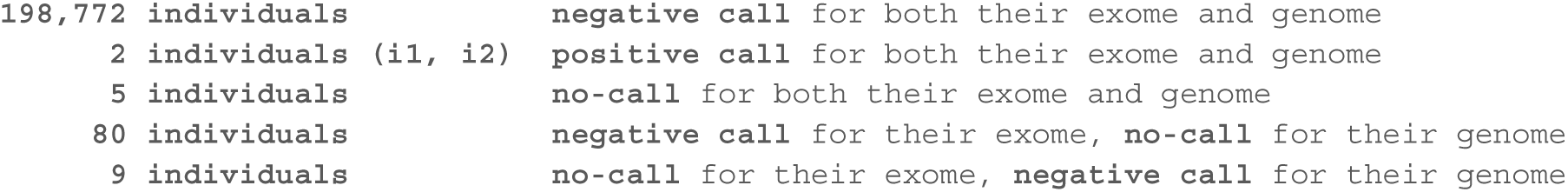
Concordance between SMA Finder calls in UKBB exomes vs genomes.

### SMN sequences within GRCh38 ALT contigs

In addition to the standard *SMN1* and *SMN2* gene annotations on chr5, the GRCh38 reference contains three more copies of SMN within its ALT contigs. Specifically, a copy of *SMN1* can be found at chr5_KI270897v1_alt:473491-501447, a copy of *SMN2* occurs at chr5_KI270897v1_alt:274951-303848, and a reverse complement sequence of *SMN1* occurs at chr5_GL339449v2_alt:456849-485731. Hypothetically, during read alignment, these extra copies could absorb informative reads and distort the chr5 read counts that SMA Finder relies on to distinguish positive from negative samples. In practice, the BWA MEM aligner performs ALT-aware alignment so that, if a read has comparable alignments to both chromosomal and ALT contig sequence(s), BWA will report the chromosome alignment as the primary one, while any ALT contig alignments will be marked as secondary ^†^. Since SMA Finder does not count secondary alignments, we assumed that it would be safe to ignore these extra SMN sequences on ALT contigs. To test the validity of this assumption, we reanalyzed all exome and genome samples within the CMG cohort to confirm that there were no primary alignments overlapping the c.840 positions of these three extra SMN copies on ALT contigs. Specifically, we used a modified version SMA Finder to count reads at the following genomic positions which correspond to c.840 of the three alternative SMN sequences: chr5_KI270897v1_alt:500378 (*SMN1*), chr5_KI270897v1_alt:301867 (*SMN2*), and chr5_GL339449v2_alt:458845 (reverse complement *SMN1*). As expected, in all CMG samples there were zero primary alignments overlapping these three positions, confirming that SMA Finder could ignore the GRCh38 ALT contigs as long as its input read data was aligned using BWA MEM or a functionally-equivalent aligner.

† See https://github.com/lh3/bwa/blob/master/README-alt.md#step-1-bwa-mem-mapping for a detailed description of BWA’s approach to ALT contigs.

### Detailed description of the SMA Finder algorithm

SMA Finder starts by retrieving aligned reads that overlap the c.840 position in *SMN1* and *SMN2*. Then, it computes two read counts, **r** and **N**, which are defined as follows:

**N** = total number of reads that overlap the c.840 position in *SMN1* + *SMN2*

**r** = the number of reads that have a ‘C’ base at the c.840 position of *SMN1* or *SMN2* and therefore support the presence of at least one intact copy of *SMN1*

These counts include reads whose mapping quality equals zero, but exclude reads whose base quality score at the c.840 position is < 13.

When **N** ≥ 14, SMA Finder uses maximum likelihood estimation to determine whether, given these counts, it is more likely that the sample has zero functional copies of *SMN1*, or that it has more than zero copies. Otherwise, if **N** < 14, SMA Finder reports that the sample has insufficient read coverage to make a call.

When computing likelihoods, SMA Finder makes the following assumptions:

**Assumption 1:** The probability that a sequencing error caused a particular base within a given read to be incorrectly called a ‘T’ when the true base was a ‘C’ is = 0.001 / 3. This is similar to calculations used within GATK HaplotypeCaller’s reference confidence model.^7^ We do not rely on base quality scores for this parameter since their accuracy can depend on the sequencing technology used and whether algorithms like GATK Base Quality Score Recalibration were applied prior to running SMA Finder. Instead, we uniformly set **p_error = 3.3e-4**.

**Assumption 2:** Let’s say **SMN1_cn** and **SMN2_cn** represent the copy numbers of *SMN1* and *SMN2* in a given sample. If an individual has at least one functional copy of *SMN1*, then the probability of the observed read counts **r** and **N** can be modeled by the binomial distribution B(**r, N, p)** where **p** = **SMN1_cn** / (**SMN1_cn** + **SMN2_cn**). On the other hand, if an individual has zero intact *SMN1* copies due to a mutation disrupting the *SMN1* c.840 position, any observed reads with a C at that position are assumed to be sequencing errors, and so the read counts are instead modeled by B(**r, N, p_error**).

**Assumption 3:** We assume that, in any given sample, the true value of **SMN1_cn** + SMN2_cn ≤ **5** based on the data in Chen et al.^3^ which found that genomes rarely contain more than 5 total copies of these paralogs.

Given these assumptions, the algorithm computes the likelihood of observing counts **r** and **N** in each of six scenarios: **SMN1_cn = 0, 1, 2, 3, 4,** or **5**, while keeping **SMN1_cn + SMN2_cn = 5**. Then, it concludes that a sample has zero functional copies of *SMN1* if the likelihood for **SMN1_cn** = 0 is greater than the likelihood for any other value of **SMN1_cn** between 1 and 5.

Since we don’t know the true value of **SMN1_cn + CMN2_cn** in a given sample, we always set **SMN1_cn + SMN2_cn** to 5 when computing likelihoods because the resulting model is the least likely to produce a false positive result compared to if we had chosen a smaller value for **SMN1_cn + SMN2_cn**.

SMA Finder also computes a Phred-scaled confidence score by subtracting the log likelihood for **SMN1_cn** = 0 from the maximum log likelihood for **SMN1_cn** = i when i is between 1 and 5: **confidence score** = 10 * abs(log10(B(r, N | **SMN1_cn = 0** )) - log10(max( { B(r, N | **SMN1_cn = i**) : i = 1, 2, 3, 4, 5 } ))).

## Notes

### Author Declarations

IRB of Mass General Brigham gave ethical approval for this work

### Summary of Updates

- Added co-authors involved in referring and following up on patients. - Updated Table 1 to include recently received results from genetic confirmatory testing of previously-diagnosed cases. - Added brief section to the discussion about estimating the population frequency of unaffected individuals with biallelic loss of SMN1.

